# Hyperacusis correlates with active and chronic-recurrent Vogt-Koyanagi-Harada disease

**DOI:** 10.1101/2025.07.17.25331731

**Authors:** Cristhian A. Urzua, Fabian Vega-Tapia, Alexis Leiva, Loreto Cuitino, Paul H Delano, Juan C. Maass

## Abstract

**Importance:** Vogt-Koyanagi-Harada disease (VKH) is a severe ocular disorder with multi-organ affection including the ear. Few studies have evaluated otolaryngologic manifestations. Current classification criteria of uveitis only includes tinnitus and disacusis.

**Objective:** To describe audiological and vestibular findings of VKH in Chile, and determine correlations with disease activity.

**Design:** Cross sectional study of Chilean patients with VKH.

**Setting:** Patients assessed between 2018 and 2021 with demographics, ophthalmologic, audiological and otolithic evaluations.

**Participants:** Subjects accomplishing VKH diagnostic criteria of the International Nomenclature Committee in Uveitis recruited in a referral academic center in Santiago, Chile.

**Main Outcome and Measures:** Primary outcome was the correlation between VKH activity and neurotologic symptoms. Demographics, symptoms and signs were assessed by structured survey and physical examination. Disability was assessed by DHI and HIIE-S. Neuro-otologic diagnostics tests were cVEMP, audiogram, suprathreshold click ABR, MLAEP, and DPOAE. Complete case correlation analysis was performed.

**Results:** 54 patients were evaluated. 65% reported ear symptoms at disease presentation. 32% of ear symptoms started synchronically and 30% started before eye symptoms. Ear and vestibular symptoms produced predominantly mild disability. The history of nausea, falls or hyperacusis during presentation correlated with disease activity at study evaluation. Hyperacusis at evaluation correlated with active or chronic-recurrent VKH. Neurotologic tests revealed mild ear impairment and few correlations with eye disease.

**Conclusions:** Otolaryngology features are frequent in VKH and may precede eye disease. Hyperacusis at evaluation may indicate current or persistent ocular disease. A broader spectrum of neurotologic symptoms must be considered in current classification of uveitis.

**Key Points:** *Question:* Are there characteristic findings in the hearing and otolithic function of patients with Vogt-Koyanagi-Harada disease (VKH), and do they correlate to the eye disease activity?

*Findings:* In a cross sectional study of 54 patients with VKH, a high frequency of neuro-otologic manifestations, and some associations to activity, chronicity and recurrence of VKH were found. Importantly, some of the ear symptoms may precede VKH diagnosis.

*Meaning:* VKH has neurotologic features that can aid diagnosis and staging of the disease. Thus, physicians must be aware of these manifestations.

## Introduction

Vogt-Koyanagi-Harada disease (VKH) is a multisystemic inflammatory condition affecting the uvea tissue in the eyes, but also the central nervous system, the integumentary system and the ear. Ear manifestations can be previous, synchronic or posterior to eye involvement (Nogushi et al, 2014). Their inclusion into differential diagnoses of ear conditions may lead to prompt diagnosis and avoid sequelae. Despite its importance, there are relatively few studies on the neurotologic features of VKH (Morita et al., 2014).

Although the literature recommends a detailed anamnesis, few neurotologic symptoms have been reported. The ear evaluation in VKH has been mainly focussed on hearing loss, tinnitus, dizziness and vertigo (Morita et al., 2014; Fujiwara et al., 2017; Al Dousary et al, 2011; Rodriguez-Rivera et al, 2011). Ondrey and Kitamura added dysacusis to the spectrum of symptoms (Ondrey et al., 2006; Kitamura et al., 2005). Although the current classification criteria of the international committee in uveitis have been recently revised, only tinnitus and dysacusis were included as neurotologic findings (SUN Working Group, 2021).

VKH affects predominantly pigmented populations (Ondrey et al., 2006). The incidence of VKH increases in Asian countries, and in some areas of South America, such as in Chile and Argentina. However, most previous South American studies have not focused on ear manifestations (Liberman et al., 2015; Giordano et al., 2017; Guayacan et al., 2018; Marquezan MC et al., 2020). Only one retrospective study reported correlation between tinnitus and response to steroid treatment (Urzua et al., 2015). In order to enhance the awareness, it is important to have a more comprehensive and locally based description of neurotologic symptoms. It is possible that in a more extensive and ear oriented study, other symptoms or neurotology lab testing would correlate to VKH features. Therefore, taking advantage of a large cohort of Chilean VKH patients (Urzua et al., 2015) we designed a cross sectional study to evaluate neurotologic findings and their association to activity of VKH.

## Materials and Methods

### General design

A cross sectional study was conducted to describe ear symptoms, hearing and otolithic function in a group of Chilean VKH patients and to seek for associations with activity of ocular disease.

### Participants and study size

Individuals accomplishing diagnostic criteria of VKH (International Nomenclature Committee in Uveitis) (SUN Working Group, 2021) were included. Recruitment was done through convenience: all patients at the uveitic clinic from Hospital Clínico de la Universidad de Chile were invited to participate. Assessment was in two day sessions between 2018 and 2021, including epidemiological, ophthalmologic, otolaryngologic, audiological and otolithic evaluations.

### Variables, sources of data, and control of bias

Variables were defined prior to the data collection. Eye disease activity and its correlation with ear symptoms was our primary outcome.

Demographic and clinical data were recorded. General health, ocular and neurotologic symptoms present at evaluation and during VKH presentation were assessed by a structured questionnaire. The standard of care treatment received by the participants was recorded. At evaluation, quality of life associated with imbalance and hearing loss were assessed by DHI and HHIE-S surveys.

A comprehensive ocular evaluation was performed, including visual acuity, intraocular pressure, slit lamp examination, and posterior segment exploration under mydriasis, as well as ancillary tests, including Optical Coherence Tomography (OCT) and fundus fluorescein angiography when necessary. Active VKH was arbitrarily defined as the presence of inflammatory findings: chamber cells, vitreous haze equal or more than 0.5+ (Jabs et al., 2005), and subretinal fluid on fundus examination or OCT. Ocular and integumentary depigmentation findings (i.e., sunset glow fundus, Dallen-Fuchs like nodules, poliosis and vitiligo) findings were considered signs of chronic-recurrent VKH (Urzua et al., 2022).

Hearing and balance evaluations were assessed in a sound attenuating room by an expert audiologist. The absence of middle ear disease was ruled out by otoscopy and tympanometry (MT10 tympanometer Interacoustics, Middlefart, DK). Briefly, the measures performed were: audiogram with hearing thresholds determination between 125 to 16000 Hz, and loudness discomfort level (LDL) between 250 to 4000 Hz by a two-channel AC-40 audiometer (Interacoustics) and DD45 headphones (RadioEar); cVEMP (P1 and N1 amplitude, latency and asymmetry) at 100, 95 and 95 dB nHL, suprathreshold click Auditory Brainstem Response (ABR) and Middle Latency Auditory Evoked Potentials (MLAEP) at 80 to 100 dBnHL (Wave I, Wave V, P1, N1 and P2 latency and amplitude) measured in Eclipse platform with research module (EP25-Interacoustics®, Denmark); and DPgram - Distortion Product Otoacoustic Emissions (DPOAE) determination at eight frequencies between 707 and 3563 Hz (ER10C, Etymotic Research®) after stimulation at 55 and 65 dBSPL. The Hearing Field was determined as the difference between hearing thresholds and LDL from 250 to 4000 Hz. cVEMP were considered asymmetrical, when amplitude ratio (VAR) was bigger than .45.

The hearing of the subjects was classified according to the .5-2 kHz and the .5-4 kHz pure tone averages (PTA) using the WHO classification (Olusanya et al,. 2019) into normal hearing (25 dB HL or less), mild (between 26 and 40 dB HL), moderate (between 41 and 70 dB HL), and severe hearing loss (between 71 and 90 dB HL). PTA asymmetry was defined as more than 15dBHL of interaural PTA difference using the same criteria that Morita et al. (2014).

In some analyses, time from eye to ear symptoms were used to control the symptoms not related to VKH.

### Statistics

Complete case analysis was used. Normality, homoscedasticity, and type of variables were assessed. Logistic regression, T-Student, Mann-Whitney and Chi square Pearson and Fisher exact test were used to seek for associations and compare means, medians and proportions respectively. P-value ≤ 0.05 was considered significant. One tail test was used except when specified. Stata 15.1 MP (StataCorp, College Station, Tx) and R were used for statistical analysis.

### Ethics

The study was approved by the Institutional Review Board (protocol number 1019/19). All participants voluntarily signed the informed consent.

## Results

54 patients were included (51 females and 3 males). The study evaluation was performed 16 months distant from the diagnosis (median; IQR 4 - 97). All participants completed the quality of life surveys, ophthalmic, and neurotologic evaluation. 52 completed the VEMP, and 39 the DPOAE. VKH disease was considered ophthalmologically active in 19 patients at study evaluation. Table 1 shows the characteristics of the participants. Within the individuals that declared noise exposure: five declared exclusively occupational exposure, twelve recreational, and seven to both sources of noise.

**Table 1:** Demographics and clinical characteristics of participants.

|  |  | Mean±SD / Median ± IQR /<br>Number (%) | N |
| --- | --- | --- | --- |
| Demographics | Average age at VKH presentation | 33.55 ± 12.28 | 53 |
|  | Sex or Gender (Female) | 51 (94.44 %) | 54 |
|  | Native American Ethnicity (Mapuche) | 9 (16.67 %) | 54 |
|  | School and Other Studies years | 15.76 ± 3.70 | 54 |
|  | Noise exposure | 26 (48.15 %) | 54 |
|  | Ototoxic drugs exposure | 7 (12.96 %) | 54 |
| Clinical findings at<br>VKH presentation | Visual deficit | 51 (94.44 %) | 54 |
|  | Eye pain | 49 (90.74 %) | 54 |
|  | Red eye | 47 (90.38 %) | 52 |
|  | Entopsia | 36 (69.23 %) | 52 |
|  | Photopsia | 39 (75.00 %) | 54 |
|  | VKH eye activity signs (any) | 19 (35.19 %) | 54 |
|  | Headache | 47 (87.04 %) | 54 |
|  | Nausea | 29 (56.86 %) | 51 |
|  | Vomiting | 17 (32.08 %) | 53 |
|  | Head paresthesia | 22 (44.00 %) | 50 |
|  | Neck stiffness | 35 (68.63 %) | 51 |
|  | Baldness | 25 (48.08 %) | 52 |
|  | Gray hair (achromotrichia) | 37 (68.52 %) | 54 |
|  | Vitiligo | 15 (27.78 %) | 54 |
|  | Time from VKH presentation to study<br>evaluation (months) | 53.98 ± 72.32 | 53 |
| Neuro-otologic<br>symptoms at VKH<br>presentation | Auditory System Symptoms (any) | 36 (67.92 %) | 53 |
|  | -Tinnitus | 30 (56.60 %) | 53 |
|  | -Deafness | 14 (27.45 %) | 51 |
|  | -Aural Plenitud | 26 (50.98 %) | 51 |
|  | -Hyperacusis | 17 (34.00 %) | 50 |
|  | -Dysacusis (paracusis, diplacusis) | 13 (25.00 %) | 52 |
|  | -Other Ear Symptoms | 13 (24.07 %) | 54 |
|  | Otalgia-ear pain | 12 (22.22 %) | 54 |
|  | Bubbles exploding | 1 (1.85 %) | 54 |
|  | Time from VKH presentation to ear symptoms (median days) | -71.95 ± 616.61 | 42 |
|  | Vestibular System Symptoms (any) | 35 (64.81 %) | 54 |
|  | -Dizziness | 32 (59.26 %) | 54 |
|  | -Unsteadiness | 29 (53.70 %) | 54 |
|  | -Vertigo | 21 (38.89 %) | 54 |
|  | -Oscillopsia | 16 (31.37 %) | 51 |
|  | -Lateropulsion | 25 (46.30 %) | 54 |
|  | -Falls | 9 (16.67 %) | 54 |
|  | -Other vestibular symptoms | 8 (15.09 %) | 53 |
|  | Cinetosis | 3 (5.56 %) | 54 |
|  | Distortion in height perception | 4 (7.41 %) | 54 |
|  | Visual vertigo | 2 (3.70 %) | 54 |
|  | Time from VKH presentation to vestibular symptoms (median days) | -196.39 ± 666.42 | 38 |

Most individuals reported neuro-otologic symptoms (Table 1 and 2). 32.08 % of patients presented auditory symptoms before the onset of eye symptoms, 30.19 % had them synchronically, and 37.74 % posteriorly (with data available; n = 53). Most auditory symptoms concentrated and presented within a 6 week period centered at the onset of eye symptoms (77.7 % of individuals). In this group, mean time from eye to auditory symptoms was -3.2 days (± 6.39 SD, median 0 days), (Figure 1). 48.12 % of individuals reported that auditory symptoms progressed after installation. The progression was from days to years with a bimodal peak (days and months were more frequent than weeks and years). 75.47 % of patients were evaluated at this study more than 3 months after the VKH presentation, and 75 % of patients with auditory symptoms at presentation still had symptoms. Vestibular symptoms presentation also concentrated within the six weeks period, affecting 65.71 % of cases. In the same period the mean time from eye to vestibular symptoms start was -0.54 days (± 9.22 SD, median 0 days), (Figure 1). Vestibular symptoms progressed in 37.04 %. The vestibular symptoms were reported as continuous in 35.13 % and fluctuant in 64.86 %. The most common temporal profile of vertigo reported was recurrent with short crisis (62.16 %), followed by acute vertigo (27.03 %), recurrent with long crisis (8.11 %) and less often chronic (2.7 %).

**Figure 1:**
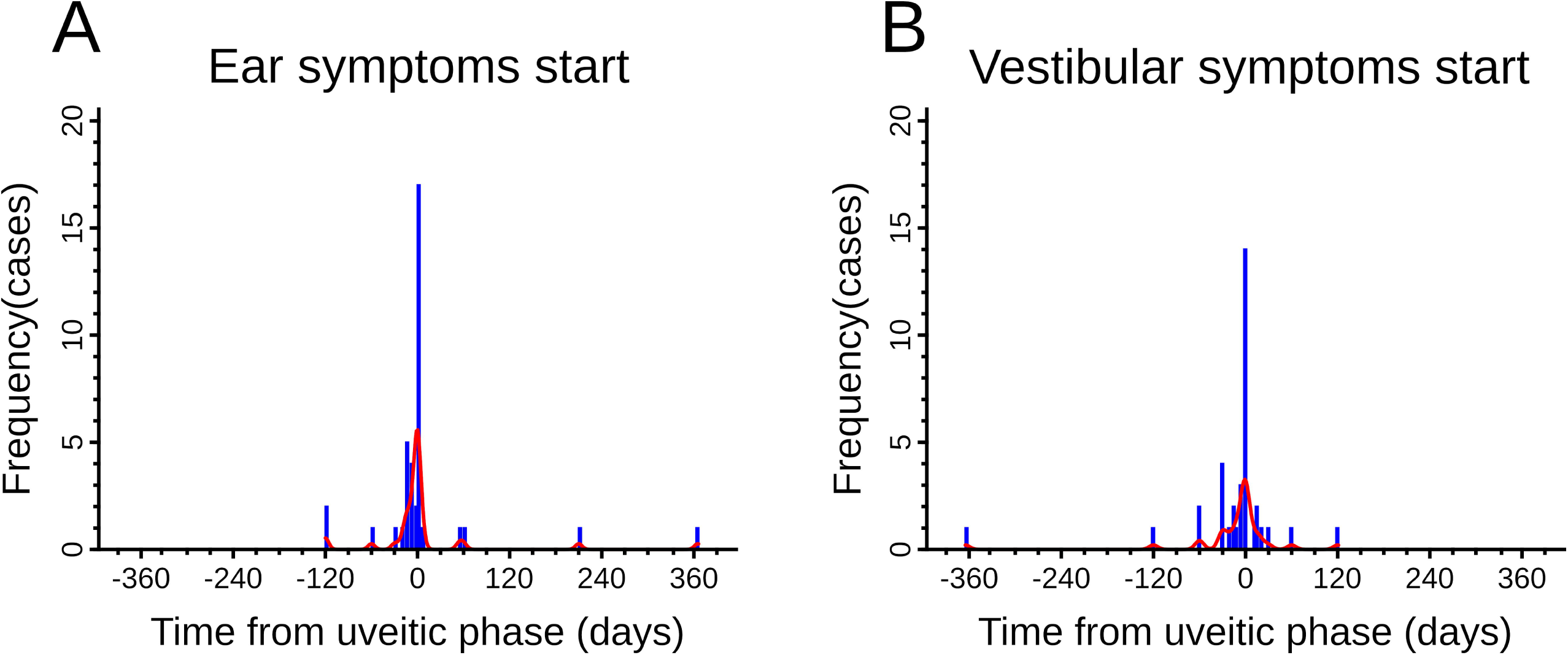
Time from eye to Neurotologic symptoms. A and B are histograms showing the number of cases presenting the symptoms and time from eye symptoms presentation to the appearance of ear (A) and vestibular (B) symptoms. Blue bars show the number of cases aggregated in 3 days bins, data treated as continuous, red lines show Kernel density plot made with Gaussian function. Minor ticks represent 30 day periods. Only individuals presenting symptoms within a 2 years period around uveitic initiation are shown. n=39 and 36 for ear and vestibular symptoms respectively.

To investigate the handicap produced by ear manifestations, HHIE-S and DHI surveys were applied. To decrease the effect of other etiologies involved in ear symptoms, only the individuals with ear symptoms that started within the six weeks period around the VKH presentation were assessed. In this group, the percentage of patients with auditory symptoms that had no significant disability was 53.33 %, 43.33 % had mild to moderate and 3.33 % had severe disability (n=30). The median HHIE-S score was 8, ranging from 0 to 30 (2-14 IQR). At the DHI survey, according to Whithney et al, (2004) classification, the percentage of patients with vestibular symptoms with no significant disability was 0 %, mild 54.17 %, moderate 29.17 % and 16.67 % severe (n = 24). The median DHI score in this group was 29, ranging from 2 to 86 (19 - 53 IQR).

Ear diagnostic tests showed mild alterations (Figure 2, Tables 1B and 2). Hearing thresholds shifts (0.125 to 16 KHz) were present in 44 patients out of 54. Most of them were bilateral and symmetrical. When ear symptoms started six weeks around disease presentation, the proportion of shifts was 80 % when having symptoms at evaluation and 25 % when they did not. In general, the rest of measurements, including PTA, LDL, discrimination scores, DPOAE, ABR, MLAEP, and cVEMP were within the normal range and were altered in very few individuals. Only five individuals out of 54 presented hearing loss according to PTA 0.5-4 KHz: three of them were unilateral and two were bilateral, both symmetrical. The hearing loss degree was found mild in three individuals, while moderate and severe in only one case each. Only three individuals out of 39 tested did not present DPOAE in both ears and one in just one ear.

**Figure 2:**
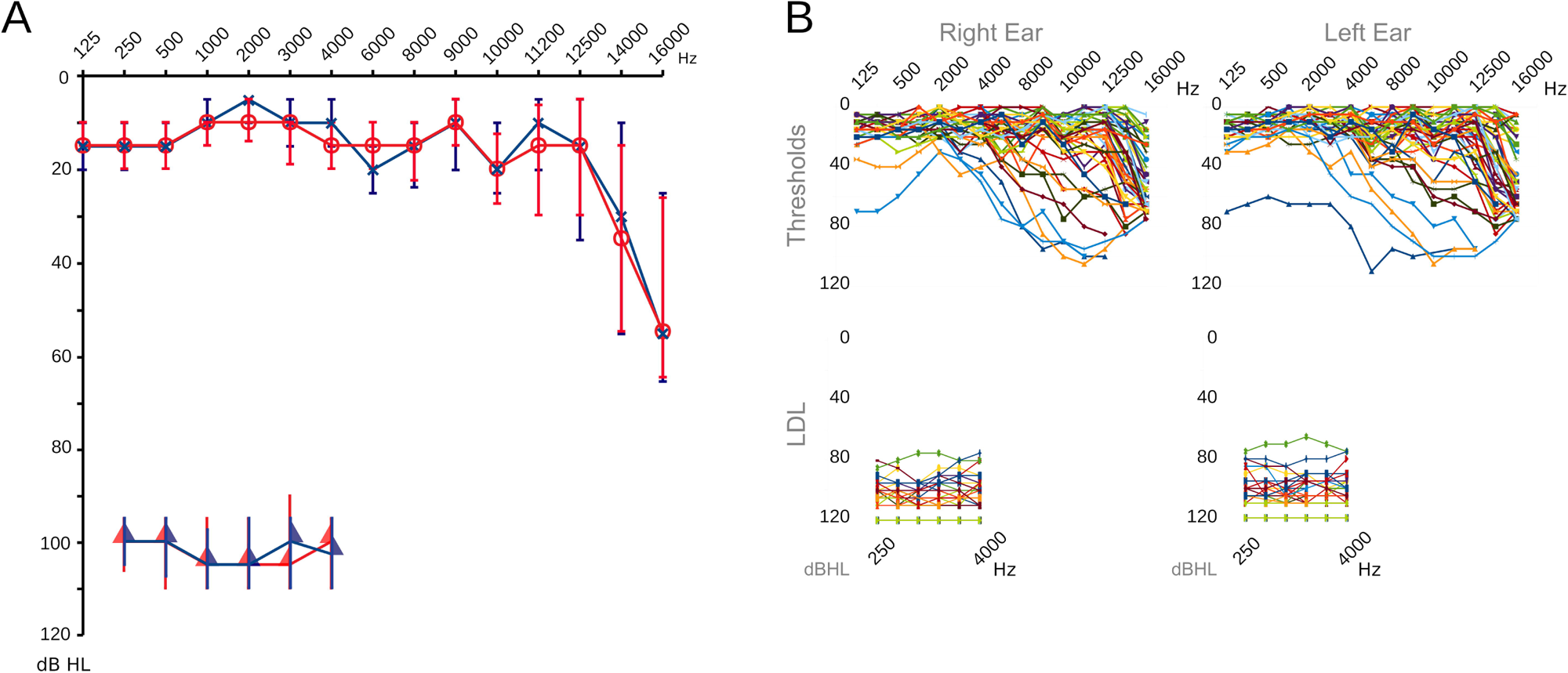
Summary audiogram of the cohort. A.- Median hearing thresholds and Loudness Discomfort Level (LDL) of the Cohort. Hearing thresholds are represented by circles (right ear) and crosses (left ear). Rectangle triangles are the LDL. Frequencies in Hertz (Hz) and amplitudes in decibels (dB). The left ear is represented in blue and the right ear in red. Bars are the IQR. B.- Actual thresholds and LDL across the whole cohort. Hearing thresholds were assessed from 125 to 16,000 Hz and LDL between 250 and 4,000 Hz.

To assess whether the ear manifestations and test results could be related to signs of active and chronic-recurrent eye disease, correlations were explored. Few associations were found between neurotologic evaluation and eye signs at ocular study evaluation (Figure 3 and Table 2). Nauseas, falls and hyperacusis at presentation correlated with VKH activity at evaluation (Table 2). Within hearing or vestibular tests at evaluation, only some hearing thresholds significantly correlated to acute inflammation signs (Figure 3). Interestingly, after adding signs of recurrent disease (sunset glow fondus or vitiligo) to signs of active disease, we found that hyperacusis at evaluation (26 vs 57%, Pearson Chi2 p-value = 0.009, n = 54) and reduction of hearing field at 500 Hz at evaluation (90 vs 80 dBHL Mann Whitney p-value = 0.0348, n = 29) positively correlated with signs of “active or chronic-recurrent” eye disease at evaluation. Finally, we generated two logistics regression models: one to predict activity and other to predict either activity or recurrence at evaluation using current and previous symptoms and antecedents. The models predicted “disease activity” with Chi2 p-value of 0.0023 and a pseudo R2 of .65 (using age, nausea, falls and hyperacusis at presentation and hearing fields at evaluation) and predicted “active or chronic-recurrent disease” with a Chi2 p-value of 0.0011 and pseudo R2 of .70 (using age, nausea and falls at presentation and hyperacusis and hearing fields at evaluation).

**Figure 3.**
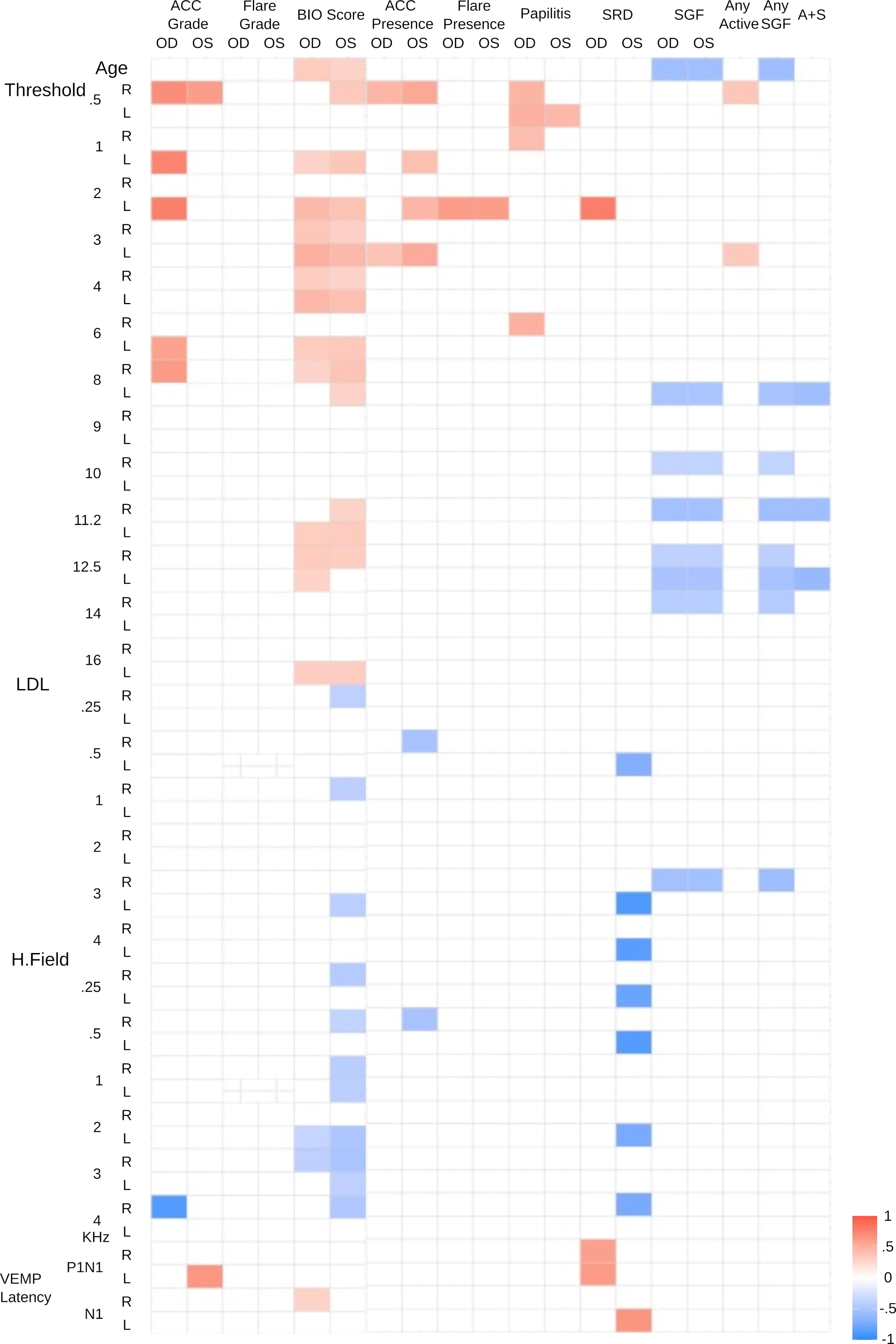
Ear and Eye features correlations. Heatmap representing correlations between Neurotology tests and Eye features. Rows show correlations with hearing thresholds between .5 to 16 KHz, LDL (loudness discomfort level) between .5 to 4 KHz, hearing field between .5 to 4 KHz and P1N1 and N1 VEMP Latencies for the left (L), and the right (R) ear. Columns show ACC (Anterior Chamber Cells) grade, Flare grade, BIO (Binocular Indirect Ophthalmoscopy) score, ACC presence, Flare presence, Papillitis, SRD (Serous Retinal Detachment) and SGF (Sunset Glow Fundus) for the right (OD) and the left (OS) eye. In the six first left columns, quantitative versus quantitative variables were analyzed by using Kendall correlation. In the next 13 columns to the right, the correlation was evaluated between quantitative versus qualitative variables using rank-biserial correlation (Rrb) from Mann Whitney test. All two tails test. Only significant correlations p<0.05 are shown. The color code shows correlation strength, blue for negative and red for positive correlations. A+S: Any activity sign or sunset glow fundus.

**Table 2:**
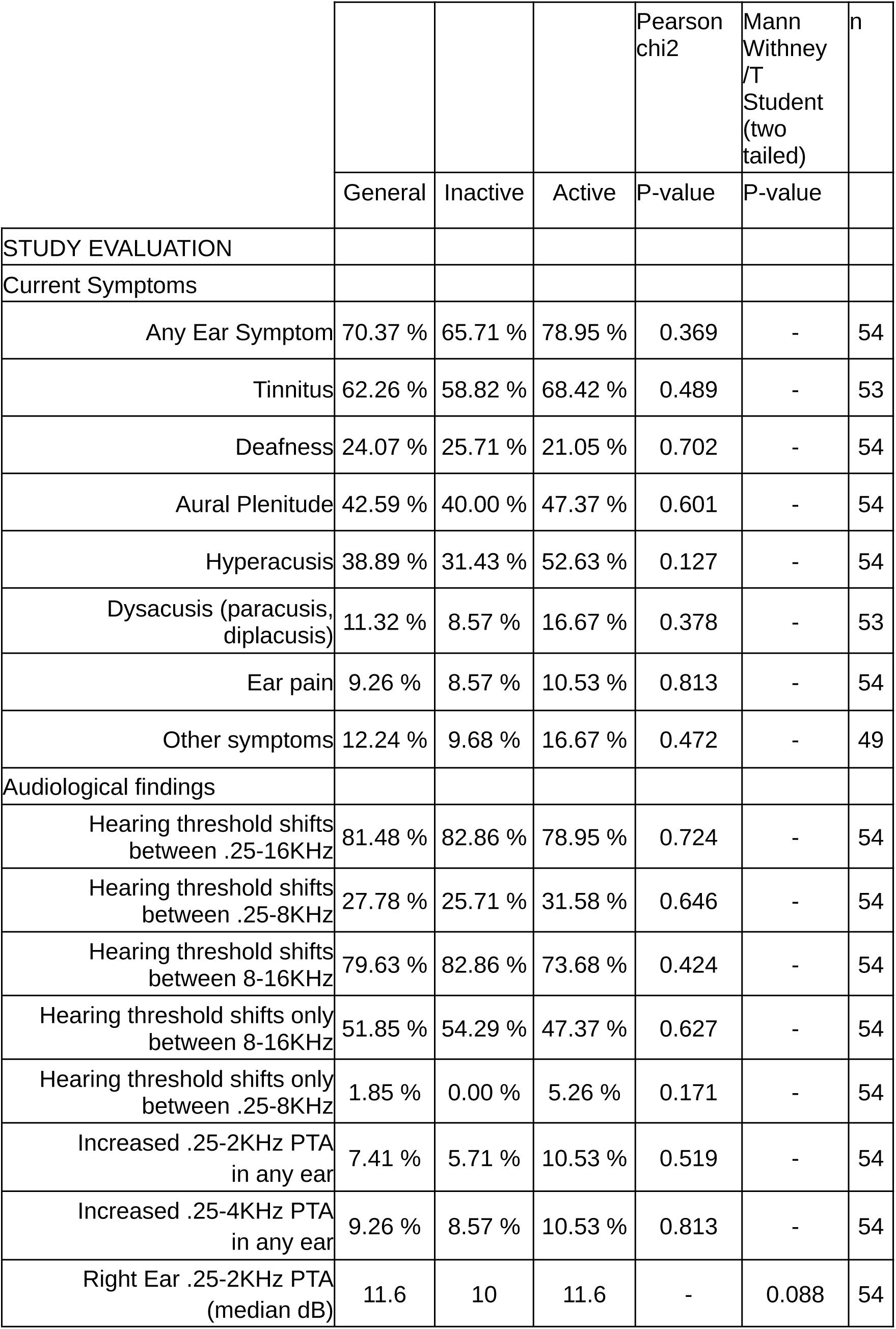
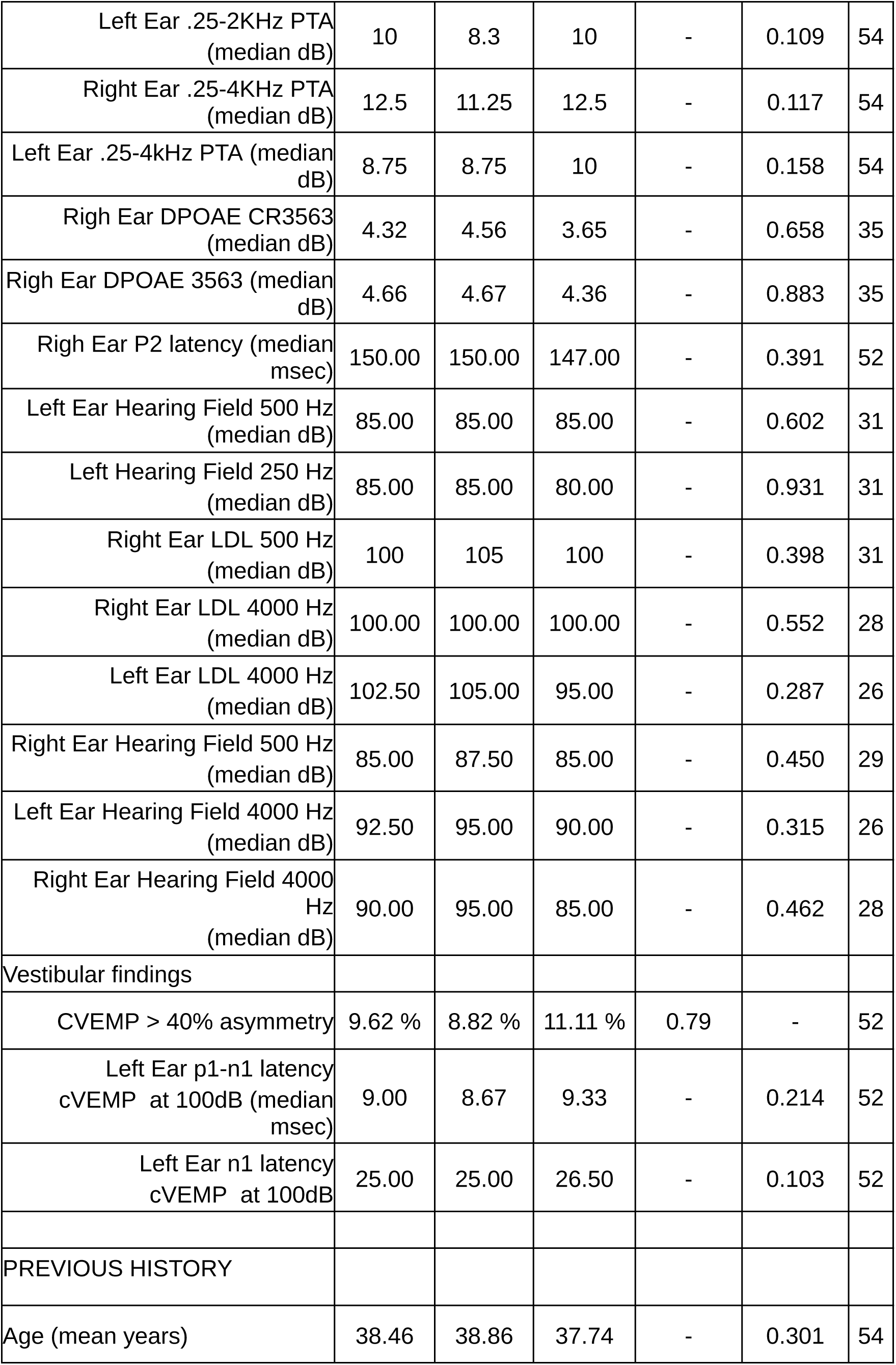
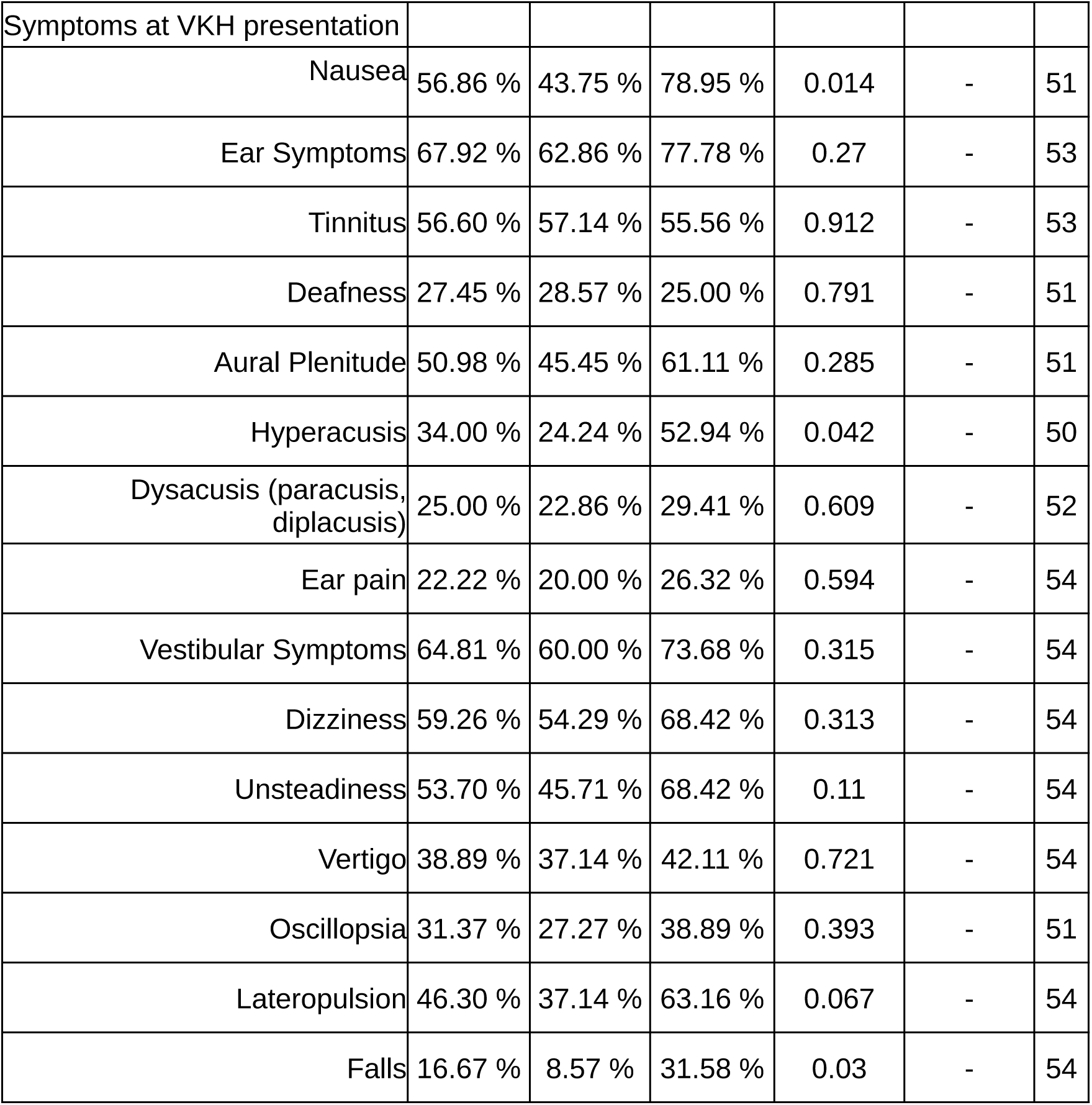
Frequency of Audiologic and Vestibular findings and correlation to VKH activity.

|  |  |  |  | Pearson<br>chi2 | Mann<br>Withney<br>/T<br>Student<br>(two<br>tailed) | n |
| --- | --- | --- | --- | --- | --- | --- |
|  | General | Inactive | Active | P-value | P-value |  |
| STUDY EVALUATION |  |  |  |  |  |  |
| Current Symptoms |  |  |  |  |  |  |
| Any Ear Symptom | 70.37 % | 65.71 % | 78.95 % | 0.369 | - | 54 |
| Tinnitus | 62.26 % | 58.82 % | 68.42 % | 0.489 | - | 53 |
| Deafness | 24.07 % | 25.71 % | 21.05 % | 0.702 | - | 54 |
| Aural Plenitude | 42.59 % | 40.00 % | 47.37 % | 0.601 | - | 54 |
| Hyperacusis | 38.89 % | 31.43 % | 52.63 % | 0.127 | - | 54 |
| Dysacusis (paracusis,<br>diplacusis) | 11.32 % | 8.57 % | 16.67 % | 0.378 | - | 53 |
| Ear pain | 9.26 % | 8.57 % | 10.53 % | 0.813 | - | 54 |
| Other symptoms | 12.24 % | 9.68 % | 16.67 % | 0.472 | - | 49 |
| Audiological findings |  |  |  |  |  |  |
| Hearing threshold shifts<br>between .25-16KHz | 81.48 % | 82.86 % | 78.95 % | 0.724 | - | 54 |
| Hearing threshold shifts<br>between .25-8KHz | 27.78 % | 25.71 % | 31.58 % | 0.646 | - | 54 |
| Hearing threshold shifts<br>between 8-16KHz | 79.63 % | 82.86 % | 73.68 % | 0.424 | - | 54 |
| Hearing threshold shifts only<br>between 8-16KHz | 51.85 % | 54.29 % | 47.37 % | 0.627 | - | 54 |
| Hearing threshold shifts only<br>between .25-8KHz | 1.85 % | 0.00 % | 5.26 % | 0.171 | - | 54 |
| Increased .25-2KHz PTA<br>in any ear | 7.41 % | 5.71 % | 10.53 % | 0.519 | - | 54 |
| Increased .25-4KHz PTA<br>in any ear | 9.26 % | 8.57 % | 10.53 % | 0.813 | - | 54 |
| Right Ear .25-2KHz PTA<br>(median dB) | 11.6 | 10 | 11.6 | - | 0.088 | 54 |
| Left Ear .25-2KHz PTA<br>(median dB) | 10 | 8.3 | 10 | - | 0.109 | 54 |
| Right Ear .25-4KHz PTA<br>(median dB) | 12.5 | 11.25 | 12.5 | - | 0.117 | 54 |
| Left Ear .25-4kHz PTA (median<br>dB) | 8.75 | 8.75 | 10 | - | 0.158 | 54 |
| Righ Ear DPOAE CR3563<br>(median dB) | 4.32 | 4.56 | 3.65 | - | 0.658 | 35 |
| Righ Ear DPOAE 3563 (median<br>dB) | 4.66 | 4.67 | 4.36 | - | 0.883 | 35 |
| Righ Ear P2 latency (median<br>msec) | 150.00 | 150.00 | 147.00 | - | 0.391 | 52 |
| Left Ear Hearing Field 500 Hz<br>(median dB) | 85.00 | 85.00 | 85.00 | - | 0.602 | 31 |
| Left Hearing Field 250 Hz<br>(median dB) | 85.00 | 85.00 | 80.00 | - | 0.931 | 31 |
| Right Ear LDL 500 Hz<br>(median dB) | 100 | 105 | 100 | - | 0.398 | 31 |
| Right Ear LDL 4000 Hz<br>(median dB) | 100.00 | 100.00 | 100.00 | - | 0.552 | 28 |
| Left Ear LDL 4000 Hz<br>(median dB) | 102.50 | 105.00 | 95.00 | - | 0.287 | 26 |
| Right Ear Hearing Field 500 Hz<br>(median dB) | 85.00 | 87.50 | 85.00 | - | 0.450 | 29 |
| Left Ear Hearing Field 4000 Hz<br>(median dB) | 92.50 | 95.00 | 90.00 | - | 0.315 | 26 |
| Right Ear Hearing Field 4000<br>Hz<br>(median dB) | 90.00 | 95.00 | 85.00 | - | 0.462 | 28 |
| Vestibular findings |  |  |  |  |  |  |
| CVEMP > 40% asymmetry | 9.62 % | 8.82 % | 11.11 % | 0.79 | - | 52 |
| Left Ear p1-n1 latency<br>cVEMP at 100dB (median<br>msec) | 9.00 | 8.67 | 9.33 | - | 0.214 | 52 |
| Left Ear n1 latency<br>cVEMP at 100dB | 25.00 | 25.00 | 26.50 | - | 0.103 | 52 |
| PREVIOUS HISTORY |  |  |  |  |  |  |
| Age (mean years) | 38.46 | 38.86 | 37.74 | - | 0.301 | 54 |
| Symptoms at VKH presentation |  |  |  |  |  |  |
| Nausea | 56.86 % | 43.75 % | 78.95 % | 0.014 | - | 51 |
| Ear Symptoms | 67.92 % | 62.86 % | 77.78 % | 0.27 | - | 53 |
| Tinnitus | 56.60 % | 57.14 % | 55.56 % | 0.912 | - | 53 |
| Deafness | 27.45 % | 28.57 % | 25.00 % | 0.791 | - | 51 |
| Aural Plenitude | 50.98 % | 45.45 % | 61.11 % | 0.285 | - | 51 |
| Hyperacusis | 34.00 % | 24.24 % | 52.94 % | 0.042 | - | 50 |
| Dysacusis (paracusis, diplacusis) | 25.00 % | 22.86 % | 29.41 % | 0.609 | - | 52 |
| Ear pain | 22.22 % | 20.00 % | 26.32 % | 0.594 | - | 54 |
| Vestibular Symptoms | 64.81 % | 60.00 % | 73.68 % | 0.315 | - | 54 |
| Dizziness | 59.26 % | 54.29 % | 68.42 % | 0.313 | - | 54 |
| Unsteadiness | 53.70 % | 45.71 % | 68.42 % | 0.11 | - | 54 |
| Vertigo | 38.89 % | 37.14 % | 42.11 % | 0.721 | - | 54 |
| Oscillopsia | 31.37 % | 27.27 % | 38.89 % | 0.393 | - | 51 |
| Lateropulsion | 46.30 % | 37.14 % | 63.16 % | 0.067 | - | 54 |
| Falls | 16.67 % | 8.57 % | 31.58 % | 0.03 | - | 54 |

## Discussion

This study found significant correlation between current activity of VKH and presence of symptoms attributed to the auditory and vestibular systems at presentation. Consecutively, the isolated finding of nausea, falls or hyperacusis at presentation may suggest a potential active disease in future evaluations. Although we did not assess the possibility directly, this association perhaps reflected a longer or more severe disease. Interestingly, we found association to current hyperacusis at evaluation only when we aggregated the chronic-recurrent and acute eye VKH signs into one variable. The signs of sunset glow and vitiligo (Urzua et al., 2022) did not correlate to ear symptoms alone, but it did correlate to them when it was aggregated with active VKH signs. Previously our group found that tinnitus negatively correlated to steroid therapy response (Urzua et al., 2015) but we did not find any trend in that direction in the present study. Perhaps the timing of our study, or the high prevalence and persistence of the symptoms explained the lack of more correlations. This is the first study seeking correlations between extraocular findings with the activity of the disease.

Chronic recurrent VKH represents a distinctive stage of the disease, characterized by ocular and integumentary depigmentation findings, the presence of recurrent episodes of inflammation, refractoriness to therapy, and higher frequency of ocular complications, as well as worse functional outcomes (Urzua et al.,2022). In the present study, active intraocular inflammation, vitiligo and sunset glow fundus (two classical depigmentation signs in skin and eyes, respectively) were associated with hyperacusis. This auditory symptom is easy to be explored in a regular clinical basis, and it may help ophthalmologists to make management decisions, since its presence would be associated with active ocular inflammation and with the worse VKH stage. In that sense, detection of inflammation in VKH may be a dilemma, since some patients persist with subclinical inflammation, which is not detectable with the routine diagnostic procedures (Murata et al., 2019; Kawaguchi et al., 2010). Moreover, evidence of higher risk of chronic recurrent VKH should guide the clinician to initiate more intensive treatment in order to catch the therapeutic window of opportunity (Herbort et al., 2019). Interestingly, it has been described that a high proportion of patients with sunset glow fundus present subclinical inflammation, which is only evident with complementary not broadly available technologies, such as Laser Flare Photometry or Indocyanine green angiography (Murata et al., 2019).

Our study found a high incidence of auditory and vestibular symptoms in VKH patients (around 65 %). Most previous studies reported 12.5 to 50 % of auditory symptoms (Ondrey et al, 2006, Morita et al, 2014, Al Dousary et al, 2011). Only Nogushi et al (2014) reported a higher 68%. Vestibular symptoms were reported in 13 to 37 % of patients (Fujiwara et al., 2017, Nogushi et al., 2014). However, these previous studies evaluated only the presence of hearing loss, tinnitus, dysacusis (paracusis or diplacusis), vertigo and dizziness at presentation, while we included a wider variety of symptoms, adding hyperacusis, aural plenitude, ear pain, falls, lateropulsion, oscillopsia, and unsteadiness. It may explain the higher incidences of symptoms found. Recently, dysacusis was included in the diagnostic criteria along with tinnitus (SUN working Group, 2021). We believe that the diagnostic work-out of VKH should be enriched with a broader group of ear symptoms.

As previously described (Nogushi et al, 2014), our results support that ear symptoms can precede, be coincident or follow the ocular presentation of VKH. Which is important since dysacusis, hyperacusis, aural plenitude or dizziness with falls are not frequent otolaryngology complaints. Thus, upon these symptoms, the physician should consider the co-occurrence of eye disease.

Neurotology lab work was predominantly normal, but also showed some abnormal tests even in absence of symptoms. Most of the alterations were found in high frequency hearing thresholds. As it was similarly described by Morita et al (2014), the rate of hearing threshold shifts detected in our study was higher than the subjective report by the patients. The rate of abnormal audiograms (.125 to 8 KHz) in this study was similar to previously published by Morita et al, (2014). They reported 74.8 % of normal audiograms (.125 to 8 KHz) at later evaluation. This study did not perform evaluations at the VKH presentation. cVEMP showed alterations in few patients, but they positively correlated to hearing test alterations, as much as vestibular symptoms correlated to hearing symptoms supporting a whole inner ear damage. This is one of the few studies evaluating vestibular phenotype of VKH patients (Fujiwara et al., 2017, Nogushi et al., 2014). Even though most tests showed rare and mild to moderate alterations, physicians must evaluate hearing and balance in these patients. Finding any other than eye alterations will support prompt systemic treatment initiation and advise a closer follow-up.

This is the first study that includes quality of life associated with neurotologic symptoms in this disease. Although our study detected auditory symptoms and threshold shifts at high frequency audiograms, the handicap associated with deafness was predominantly mild. Perhaps this was because hearing loss is less notorious than blindness. Instead, the handicap associated with vestibular symptoms was more relevant. Probably due to the concomitant affection of two systems involved in balance (eye and ear). Therefore, balance support should be considered in these patients at presentation.

## Limitations

As a cross sectional study, the previous symptoms and antecedents reported in this study were obtained by surveys, thus recall bias is possible. The noticeability and severity of symptoms might be affected by the time lapse between presentation and evaluation. Despite this, many symptoms were recalled in our study. We found a frequent report of hyperacusis, or the abnormally increased and painful ear sensation after sound. A symptom not commonly recognized within the spectrum of VKH symptoms (Morita et al., 2014; Fujiwara et al., 2017; Al Dousary et al, 2011; Rodriguez-Rivera et al, 2011; Ondrey et al., 2006; Kitamura et al., 2005). Although we differentiated hyperacusis and dysacusis (paracusis, diplacusis), some groups include the term hyperacusis in dysacusis. Thus, in that last case our study would find dysacusis (only as hyperacusis) instead. Specifically, the paracusis form of dysacusis did not correlate to eye disease of VKH. It might be useful to add a specific definition of dysacusis or the inclusion of other symptoms such as hyperacusis to the VKH awareness and diagnostic criteria (SUN Working Group, 2021).

## Interpretation and generalizability

Due to the higher frequency of VKH in Chile (17 % of uveitis) and the fact that 93 % of subjects were women, this study predominantly reflects the characteristics of VKH in Chilean women.

## Future evaluations

Future studies could improve the characterization of each neurotologic symptom and its evolution. The regular inclusion of auditory and vestibular symptoms in VKH surveys may decrease recall bias and would confirm the prevalence and role of these symptoms. Further studies on this cohort of patients will unveil the long-term hearing and balance of VKH patients and will ponder the neurotologic symptoms for staging and clinical follow up.

## Conclusion and relevance statement

Ophthalmologists and Otolaryngologists must be familiar with the symptoms of VKH. Neurotologic evaluation might be of benefit for these patients. Active awareness on hyperacusis screening and periodic audiograms including loudness discomfort level (LDL) and high frequencies thresholds might have a role in the follow up and staging.

## Data Availability

All data produced in the present study are available upon reasonable request to the authors

## Acknowledge section

### Conflict of interest

Authors declare no conflict of interest.

### Funding sources

This research work was supported by Fundación Guillermo Puelma to J.M. and P.D., the National Agency for Research and Development of Chile (ANID), FONDECYT 1220607, and ANID - Basal Project AFB240002 to P.D., FONDECYT grants 1212038 to C.U., 11191215 to L.C. and 3220781 to F.V.

### Contributions

J.M. and C.U. conceived the study and wrote the paper. J.M. participated in the design, implementation, and analysis of the results. C.U. participated in the study supervision, result discussion and data analysis. F.V. participated in the analysis of the data and the result discussion. P.D. participated in the study design and supervision. L.C. participated in data collection, and in study implementation, design and supervision. A.L. implemented the study and performed the data collection. All authors approved the final version of the paper.

## Notes

### Competing Interest Statement

The authors have declared no competing interest.

### Funding Statement

This research work was supported by Fundacion Guillermo Puelma to J.M. and P.D., the National Agency for Research and Development of Chile (ANID), FONDECYT 1220607, and ANID - Basal Project AFB240002 to P.D., FONDECYT grants 1212038 to C.U., 11191215 to L.C. and 3220781 to F.V.

### Author Declarations

Comite de etica asitencial del Hospital Clinico de la Universidad de Chile, Hospital Clinico Universidad de Chile, Facultad de Medicina, Universidad de Chile, approved protocol number 1019/19

